# Adaptive behavior in response to the 2022 mpox epidemic in the Paris region

**DOI:** 10.1101/2024.10.25.24315987

**Authors:** Davide Maniscalco, Olivier Robineau, Pierre-Yves Boëlle, Mattia Mazzoli, Anne-Sophie Barret, Emilie Chazelle, Alexandra Mailles, Harold Noël, Arnaud Tarantola, Annie Velter, Laura Zanetti, Vittoria Colizza

## Abstract

The 2022 mpox outbreak saw a rapid case surge among men-who-have-sex-with-men (MSM) in previously unaffected regions, driven by heterogeneity in sexual networks. A sudden decline followed, but its drivers remain unclear as it is difficult to distinguish the roles of vaccination, herd immunity, and behavioral changes. We developed a network model of mpox transmission among MSM based on sexual behavior data and fitted it to the Paris region epidemic. We studied whether the decline was driven by post-exposure prophylaxis (PEP) vaccination, immunity among highly active MSM, or behavioral changes. Behavioral shifts were modeled as either uniform or based on individual risk factors, like sexual activity or exposure to diagnosed cases. We used the cross-sectional 2023 ERAS survey to validate findings. Behavioral changes adopted by 49% (95%CI 47-51%) of MSM regardless of individual risk factors best explained the observed epidemic decline. These changes prevented an estimated 73% (28-99%) of mpox cases in summer 2022. Findings aligned with the ERAS survey data, showing that 46% (45-48%) of MSM reduced sexual partners. On the contrary, PEP vaccination and immunity among highly active MSM were insufficient to curb the outbreak. Widespread behavioral change was the primary driver of the mpox epidemic decline in the Paris region, before preventive vaccination or immunity could affect epidemic spread. These findings highlight the importance of effective risk communication and community engagement in outbreak management. Tailored public health responses that encourage adaptive behaviors, especially as vaccination efforts ramp up, are essential for supporting affected communities.

## INTRODUCTION

The 2022 global outbreak of mpox marked an international public health crisis^1^. Endemic in Western Africa, monkeypox virus clade II rapidly spread to Western Europe, then moving through Central and Southern Europe, to North America, Latin America and the rest of the world, driven largely by international travel and extensive underdetected dissemination^2^. By July 23, 2022, with more than 23,000 cases reported in 94 countries, the World Health Organization (WHO) declared mpox a Public Health Emergency of International Concern (PHEIC). As cases surged worldwide, a major concern was the mode of transmission.

The spread of mpox in 2022 was indeed remarkably different from previous outbreaks. Unlike previous mpox outbreaks that remained geographically limited to known endemic areas, the global outbreak predominantly affected men-who-have-sex-with-men (MSM^*^) and was largely associated with sexual transmission^1^. A key factor driving the rapid dissemination was the heterogeneity of sexual networks in the MSM community. A small number of individuals with an exceptionally high number of sexual partners largely amplified the disease spread, creating conditions for sustained human-to-human transmission that led to explosive case growth^2,3^.

But the explosive rise was soon followed by a sudden decline in viral circulation. This occurred as countries were struggling to implement control efforts through vaccination and risk communication. It remains uncertain whether these measures contributed to slow down the epidemic. Indeed, the rapid infection of highly active MSM could have already led to infection-induced immunity at the population level^4,5^; or individuals may have spontaneously avoided risk in response to public health messaging^6,7^. The interplay between these factors is complex and context-specific, with timing and impact varying across countries^5,8–14^, depending on factors like the time of first importations^2^, vaccination campaign onset, and resource availability. In France, vaccine post-exposure prophylaxis (PEP) of contacts of confirmed cases began on May 27, shortly after the first confirmed case on May 19, but was initially limited to 802 doses over 45 days. Mass pre-exposure mpox vaccination (mpox PrEP, hereafter referred to as PrEP) only began after the epidemic had peaked—both nationally and in the Paris region (Île-de-France), where the peak occurred in late June (week 26, June 27-July 3). Public health communication campaigns targeting MSM intensified over this period^15^. Clarifying the roles of both public health efforts and individual actions is critical for developing more effective strategies for future outbreaks.

We developed a data-driven network model of mpox transmission to identify the drivers behind the observed decline in mpox cases among MSM during the summer 2022. The study focused on the Paris region, which reported the majority of cases in France (63% at the time of the peak). Our analysis tested three main hypotheses: the impact of PEP vaccination, the role of infection-induced immunity, and behavioral changes driven by different factors. Results were then validated with surveillance data and post-outbreak behavioral survey responses.

## METHODS

Here we outline the data, models, and analyses. Additional details are provided in the Appendix.

### Mpox surveillance data

Data on the mpox outbreak were reported to Santé Publique France^16^ (French Public health Agency) for 1,616 cases in the Paris region from May 7 to September 22, 2022 (**Figure 1a**). Age, area of residence, dates of symptoms onset and testing, smallpox vaccination status and self-identification as a MSM (91% of answering cases) were collected. In the study period from May 7 to August 31, there were 1,530 cases, with 30 onset dates imputed based on known onset-to-testing delays.

**Figure 1.**
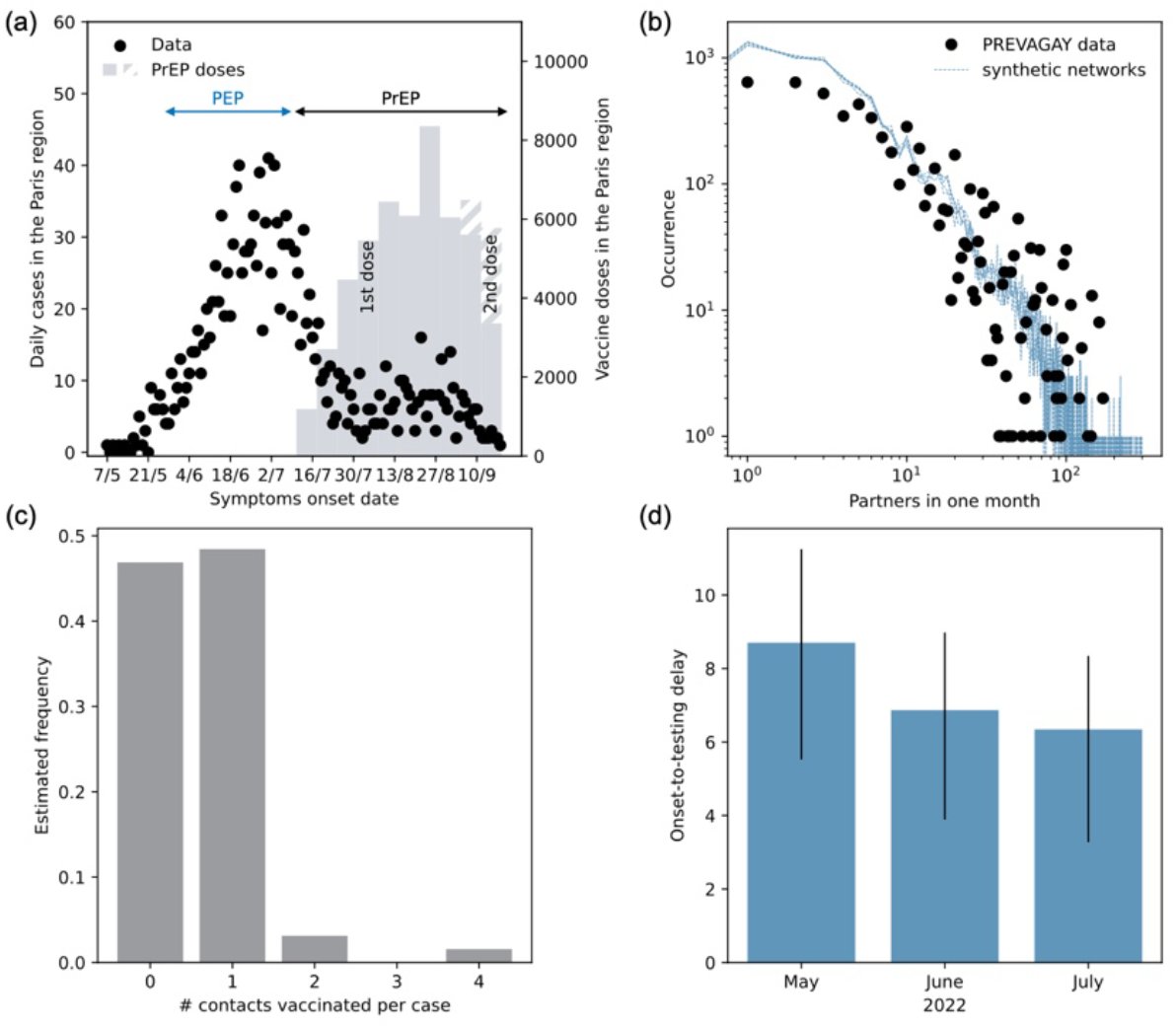
Surveillance, behavioral and vaccination data. **a:** Mpox cases in the Paris region from May 7 to September 18, 2022 (points, left y axis), together with 1^st^ (solid bars) and 2^nd^ (dashed bars) doses of PrEP vaccines in the Paris region (right y axis). **b:** Occurrence of the monthly number of sexual partners in the PREVAGAY survey and in the five synthetic networks, in log-log scale. **c:** Frequency of vaccinated contacts per mpox case estimated from contact tracing data. **d:** Onset-to-testing delay estimates by month.

### Vaccination data

Data on third-generation vaccine doses administered in the Paris region in the study period included PEP doses for at-risk contacts from May 27 to July 10, and PrEP doses starting July 11 (**Figure 1a**, see detailed timeline in **Table S2**). PEP^17^ and PrEP^18^ vaccination guidelines and the definition of at-risk contacts^19^ were defined by the Haute Autorité de Santé and Santé publique France. We also used the estimated frequency of contacts vaccinated per detected case from available contact tracing data (**Figure 1c**).

### PREVAGAY survey and MSM population estimate

The 2015 PREVAGAY survey^20,21^ included 1,089 MSM in Paris and documented age, area of residence and number of sexual partners by MSM commercial venues (bars, clubs, saunas, and others). We use these data to build a time-varying network of sexual contacts among MSM in the Paris region. The size of the MSM population in the Paris region was set to 65,000 individuals (250,000 multi-partner MSM in France^18^, 26% of whom resided in the Paris region^22^).

### ERAS 2023 survey

The ERAS survey^23^ (Enquête rapport au sexe, Survey on relation to sex) collected data from 23,502 MSM respondents across France recruited via a variety of digital media from February 24 to April 6, 2023. The survey reported data on socio-demographic characteristics, health, and sexual behavior, particularly during the 2022 mpox outbreak. Respondents included 3,965 (16.9%) multipartner MSM for the Paris region who reported whether they reduced their number of sexual partners during the outbreak.

### Ethics statement

All surveillance data were anonymised before use. Surveillance was considered as non-interventional research only requiring the non-opposition of the patient (article L1211 of the French public health code).

The PREVAGAY study was authorized by the Comité de protection des personnes Ile-de-France IX (n°2014-A01605–42). The ERAS study was approved by Inserm’s Ethics Evaluation Committee (IRB00003888 avis n°23-989). Participants in both studies gave their informed consent.

### Time-varying sexual contact network

From PREVAGAY data, we built a stochastic time-varying network of sexual contacts in MSM in the Paris region using data on age, the number of sexual partners and the commercial venues attended^20^. Five network realizations of 180 days were generated (**Figure 1b**), by sampling power law distributions fitted to reported partnership data by venue type. The PREVAGAY study and the ERAS 2023 survey reported similar figures on partnerships (**Figure S2**).

### Mpox transmission model

We described mpox transmission on the network of sexual contacts among MSM using a stochastic Susceptible-Exposed-Infected-Isolated-Recovered (SEIQR) agent-based model (**Figure S4**). Infectious individuals were identified with a detection probability *p*_*d*_ fixed to 60%^24^ (20%, 80% explored for sensitivity), except when testing the infection-induced immunity hypothesis where *p*_*d*_ was fitted. Model parameters are reported in **Table S3**. The model accounted for smallpox vaccination status and mpox vaccination with PEP and PrEP. We used 71%^25^ effectiveness for first-generation mpox vaccines (85%^26^ in the sensitivity analysis); 89%^27,28^ for third-generation Modified Vaccinia Ankara (MVA) smallpox vaccine Imvanex as PEP; and 78%^29,30^ for MVA as PrEP. Third-generation vaccines were considered effective 14 days after administration. PEP vaccination was based on the number of at-risk contacts vaccinated per detected case (**Figure 1c**), assuming only MSM unvaccinated against smallpox were eligible due to limited resources.

### Adaptive behavior models

We explored three models of behavioral changes in uninfected and unvaccinated MSM: uniform changes across the population, changes based on sexual activity, or triggered by recent exposure to diagnosed cases. Adaptive changes were modeled from mid-June to mid-July 2022, with the reduction lasting through the study period. The reduction level *r* and the percentage *c* of MSM reducing contacts were estimated by fitting each model to the epidemic decline. We tested nine periods starting on June 15, June 18, or June 21, lasting four to six weeks.

### Inference framework

We simulated a population of 10,000 MSM and rescaled it to match the Paris region population (65,000). We averaged over 250 stochastic simulations to compute the expected epidemic curve and fitted the model to observed case counts through a maximum likelihood approach. The number of fitted parameters depended on the hypothesis considered. In all cases, we fitted the rate of transmission per sexual contact (*β*) and the date of introduction of 10 seeds (*t*_*0*_). For PEP vaccination, the fit was performed on the early case rise up to June 18, for simulations with and without vaccination. For the infection-acquired immunity hypothesis, we fitted the model to the full epidemic curve and estimated *p*_*d*_ as well. For behavioral changes, we first fit *β* and *t*_*0*_ on the early rise, then estimated *c* and *r* (see above) on the remaining trajectory – assuming *β* and *t*_*0*_ were fixed. The best-fit model was determined using AIC. Simulations without change in behavior served as a counterfactual.

### Statistical analyses and validation

Monthly changes in onset-to-testing delays were tested by ANOVA. We compared mpox incidence with and without vaccination using a Chi-square test.

Uncertainty on model estimates were provided by the 95% prediction intervals (95%PI).

Model selection was validated against ERAS and WHO^7^ survey data, analyzing behavioral change responses across MSM populations in the Paris region and reported estimates for Western Europe, respectively. The reduction in the number of sexual partners obtained from ERAS data was also analyzed stratified by sexual activity. We performed a logistic regression to examine the correlation between the number of sexual partners and having changed behavior or otherwise during summer 2022 in the ERAS survey.

We compared the mean ages of infected cases over successive months (*t*-tests). Transmission heterogeneity was assessed by estimating the overdispersion parameter *k* during the epidemic rising (before June 18) and declining phases (after July 10). We compared our findings with overdispersion estimates from Paredes et al.^2^, who analyzed MPXV sequence clusters from August 2022, during the declining phase of the outbreak.

### Sensitivity analysis

We ran six sensitivity analyses, examining the effects of different detection probabilities (20%, 80%), undetected case transmissibility and vaccine protection assumptions. Behavioral change sensitivity considered both vaccinated and non-vaccinated MSM altering behavior.

## RESULTS

Based on a total of 1,500 mpox cases with complete data for the period May-August 2022, we estimated a reduction of the mean onset-to-testing delay from 8.7 (IQR 5.5-11.2) days in May to 6.9 (3.9-9.0) days in June, and 6.3 (3.3-8.3) days in July (**Figure 1d**). The decreasing trend was confirmed through an ANOVA test (p<10^-6^).

The model that best explained the observed epidemic decline assumed a uniform reduction in risk across the MSM population. It closely matched the observed epidemic curve (**Figures 2a, 2b, 3a**) and estimated that 49% (95%CI 47-51%) of MSM reduced their sexual contacts by 93% (95%CI 86-98%). This adaptation, occurring gradually between June 15 and July 20 (**Table 1**), led to a 39% (95%PI 35-41%) reduction in sexual contacts, ultimately driving the epidemic decline. By comparing the best-fit model simulations to those without behavioral changes, we found that the risk reduction prevented 73% (95%PI 28-99%) of mpox cases during the study period (**Figure 3b**).

**Table 1.** Fit results of behavioral change models.

| Model | MSM changing behavior (% of the MSM population) | Reduction in sexual activity | Avoided sexual contacts | Averted cases | Period of behavioral changes | AIC |
| --- | --- | --- | --- | --- | --- | --- |
| Model with MSM uniformly changing behavior | 49% (47-51%) | 93% (86 -98%) | 39% (35-41%) | 73% (37-98%) | 15/6 - 20/7 | 413 |
| Model with highly active MSM preferentially changing behavior | 19% (17-25%) | 96% (82-100%) | 41% (35-44%) | 73% (21-100%) | 21/6 - 26/7 | 431 |
| Model with MSM contacts of cases change behavior | 8% (0.4-13%)* | 100% (95-100%) | 28% (2-42%) | 70% (33-98%) | 21/6 - 2/8 | 452 |

\*For this model, we fitted the percentage $c$ of the contacts of mpox cases and obtained $c=100\%$ (95-100%), resulting in the percentage of the MSM population reported in the Table.

**Figure 2.**
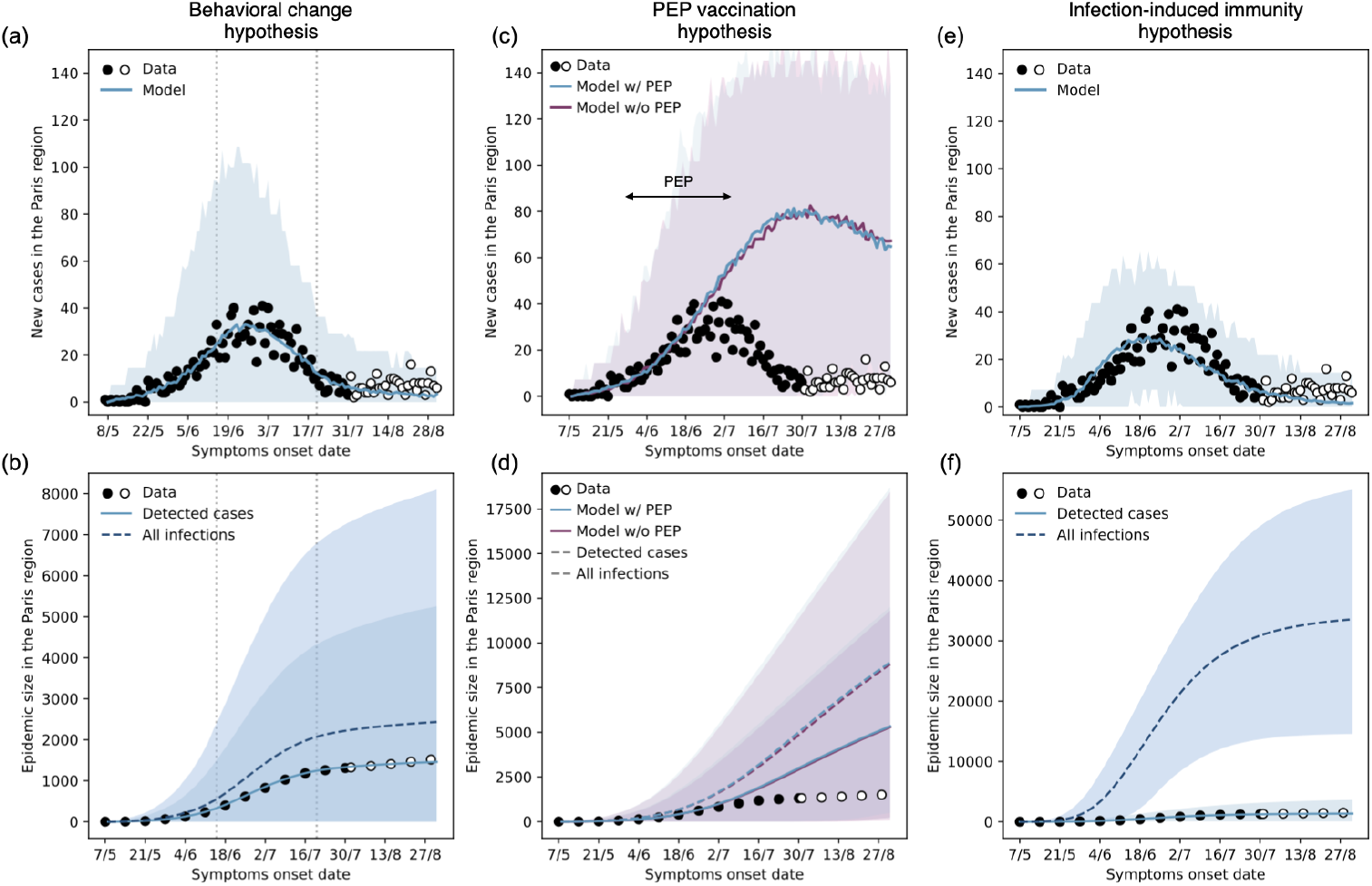
Model predictions for the three hypotheses. **a,b:** Mpox cases in the Paris region (filled points refer to the data used for inference) and model predictions with 95% prediction intervals under the behavioral change hypothesis, assuming homogeneous change of behavior across MSM (best-fit model). The top panel (a) reports incident cases, and the bottom panel (b) the epidemic size. **c,d:** As in (a,b), data vs. model predictions under the PEP vaccination hypothesis, scenarios with and without PEP vaccination. **e,f:** As in (a,b), data vs. model predictions under the infection-induced immunity hypothesis.

**Figure 3.**
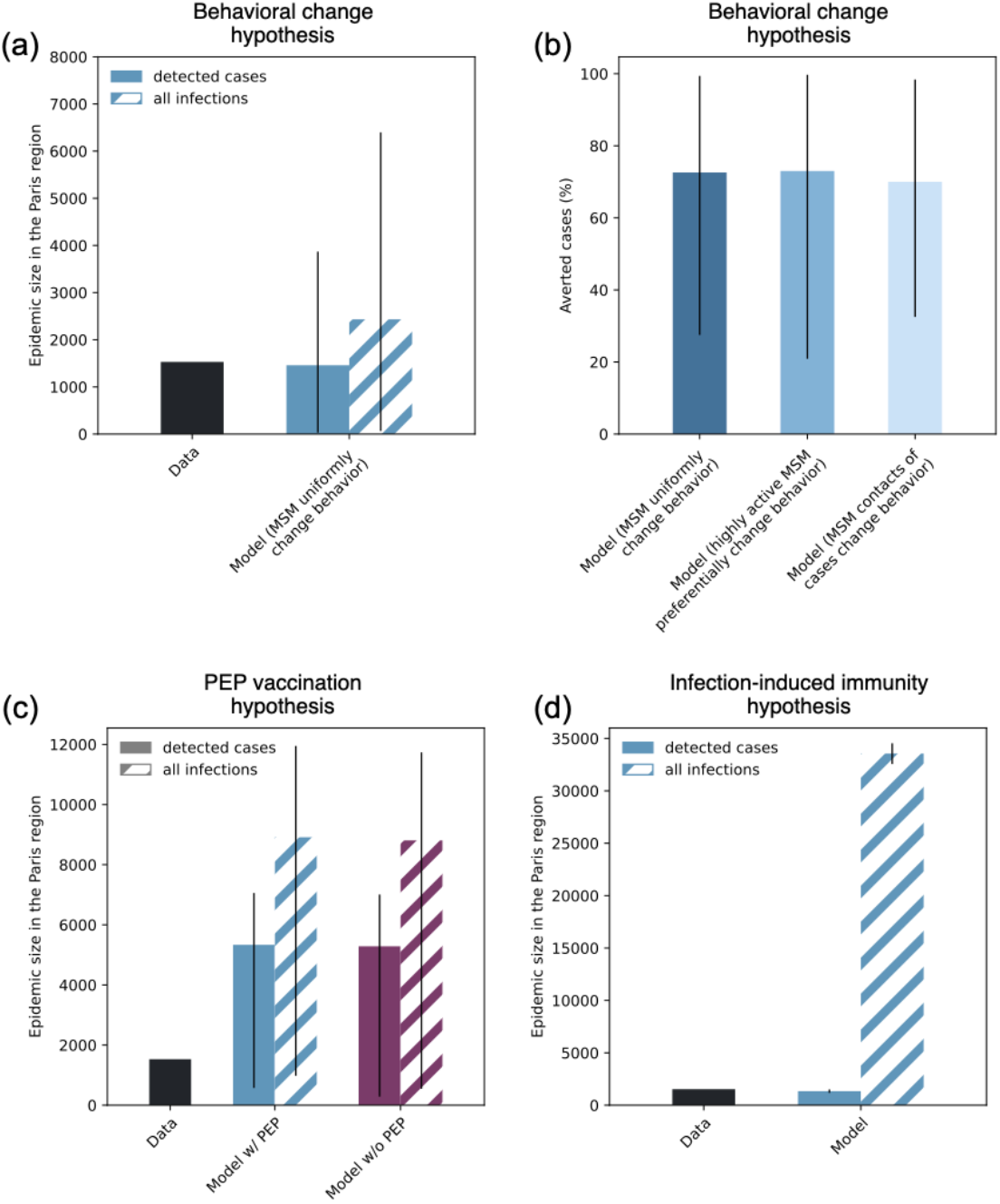
Epidemic size and averted cases. **a:** Epidemic size in the Paris region on August 31, 2022, data vs. model predictions with 95% prediction intervals in the behavioral change hypothesis, assuming homogeneous change of behavior across MSM (best-fit model). **b:** Predicted percentage with 95% prediction intervals of averted mpox cases in the Paris region from May 7 to August 31 2022 for the three assumptions considered under the behavioral change hypothesis. **c:** as in (a), data vs. model predictions with 95% prediction intervals under the PEP vaccination hypothesis, showing the two cases with and without PEP vaccination. **d:** as in (a), data vs. model predictions with 95% prediction intervals under the infection-induced immunity hypothesis.

To validate our model selection, we analyzed data from the 2023 ERAS survey, which showed that 46% (95%CI 45-48%) of MSM in the Paris region reported reducing their sexual partners during summer 2022, aligning closely with our model prediction (**Figure 4a**). Importantly, we found no significant correlation between sexual activity levels and individual risk-reduction behaviors (*p*=0.3), further supporting our model ability to capture behavioral changes across different activity levels (**Figure 4b**). In contrast, the two rejected behavioral models, which linked risk reduction to either high sexual activity or recent exposure to a detected case, did not align with observed behavioral patterns. The first model, which assumed that highly active MSM were the primary drivers of behavioral change, required only 19% (95%CI 17-25%) of MSM to reduce sexual contacts to achieve epidemic control (**Table 1**). However, this underestimated the community-wide behavioral change indicated by survey data (**Figure 4a**). The second model, which attributed behavioral changes to recent contacts of detected cases, predicted an even more modest adaptation (8%, 95%PI 0.4-13%; **Table 1**). Both models failed to reproduce the reported change of behavior of MSM with fewer than 50 partners per month (**Figures 4c, 4d**).

**Figure 4.**
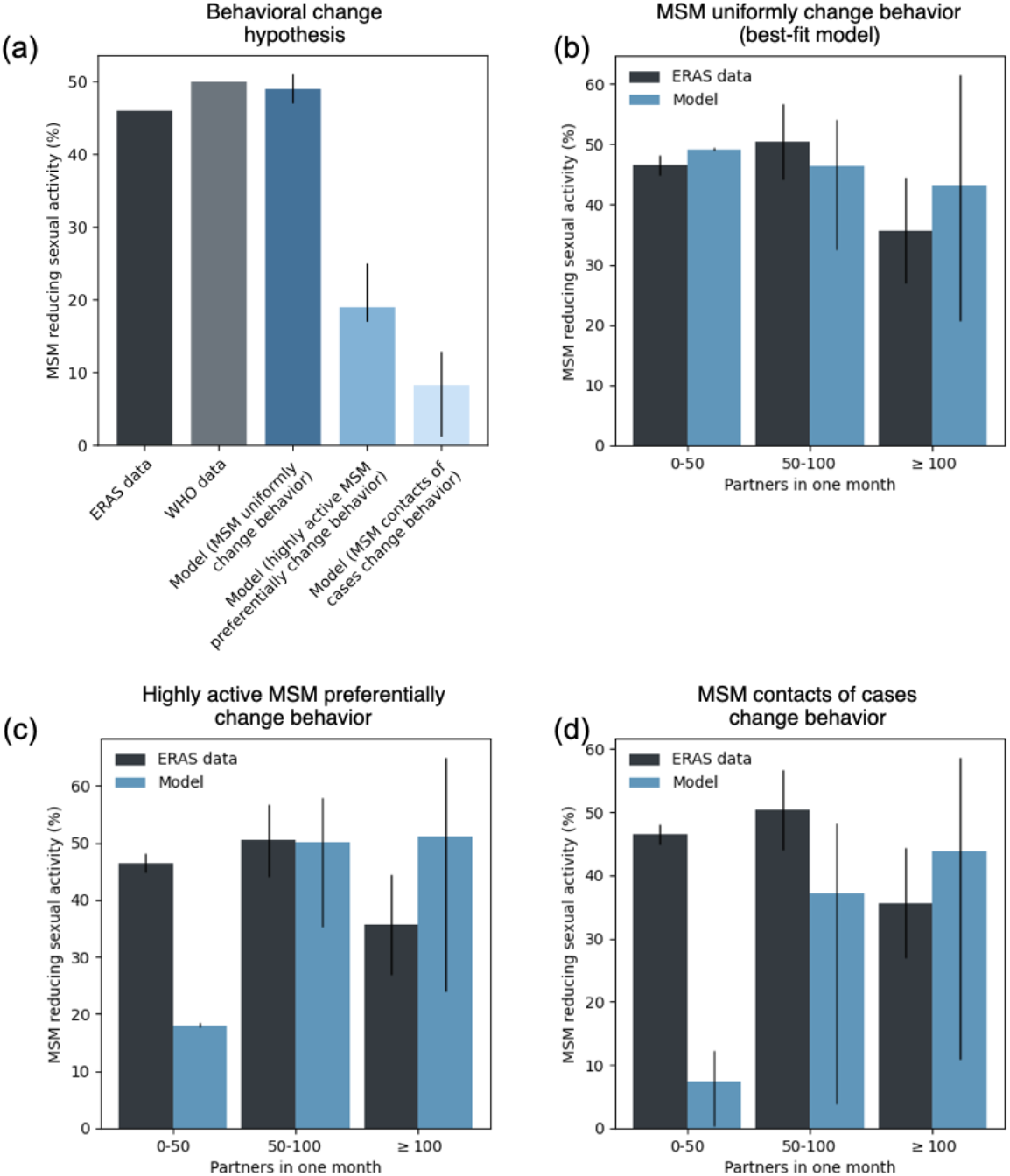
Change of behavior. **a:** Percentage of MSM reducing sexual activity: survey data vs. model estimates with 95% confidence intervals. **b:** Percentage of MSM reducing sexual activity stratified by the monthly number of partners: survey data vs. model predictions with 95% prediction intervals in the assumption of uniform change of behavior across MSM (best-fit model). **c:** as in (b) in the assumption of a change of behavior based on sexual activity. **d:** as in (b) in the assumption of a change of behavior based on recent exposure to a diagnosed case.

The infection dynamics from the best-fit model showed rapid progression, particularly among highly active MSM early in the epidemic. Despite representing only 0.4% of the MSM population, individuals with over 100 sexual contacts per month accounted for 6% (95%PI 0-13%) of early cases, declining to 4% (95%PI 0-13.5%) in the later phase (**Figure S5a**). The predicted mean age of mpox cases increased from 33 years (95%PI 20-57) in May to 38 years (95%PI 20-63) in July (t-test p < 10^−6^), aligning with surveillance data (**Figure S5b**). Simulations estimated that 25% (95%PI 5-53%) of mpox cases occurred in individuals vaccinated against smallpox, consistent with surveillance data (**Figure S5c**). Simulated transmission chains indicated substantial variation in mpox transmission, with an overdispersion parameter of 0.32 (95%PI 0.30-0.34) during the rising phase, and 0.31 (95%PI 0.29-0.34) during the decline, in agreement with estimates from sequence clusters (**Figure S5d**).

Next, we examined the role of PEP vaccination, testing whether it alone could explain the epidemic decline. Simulations with and without PEP vaccination yielded identical epidemic trajectories (**Figures 2c**), confirmed by a Chi-square test (*p*=1.0). This suggested that PEP vaccination had no significant impact on the epidemic trajectory. Most importantly, the model failed to replicate the observed incidence of cases, predicting a delayed peak by 30 days (95%PI -5-58) and a 2.7-fold higher incidence (95%PI 0.5-4.6). By August 31, the model predicted 5,335 (95%PI 573-7,058) detected mpox cases, corresponding to 8% (95%PI 0.9-11%) of the MSM population—3.5 times higher than observed (**Figures 2d, 3c**). Even a lower detection probability (*p*_*d*_=20%) – which would imply more undetected infections and higher natural immunity – could not replicate the peak timing and magnitude (**Figure S6**). We also tested whether infection-induced immunity might have curbed the epidemic due to higher circulation than actually observed, allowing for a portion of undetected cases to be estimated. While this hypothesis could fit the epidemic curve (**Figure 2e**), it required that only *p*_*d*_=4% (95%CI 4-5%) of the cases be reported. Under this scenario, 52% (95%PI 50-53%) of the MSM population in the Paris region would need to have contracted mpox by the end of August, totaling 33,561 (95%CI 32,432-34,661) cases (**Figures 2f, 3d**).

We refitted the model with uniform behavioral changes among MSM to assess the impact of different assumptions explored in the sensitivity tests. When considering a detection probability *p*_*d*_=80%, the model produced similar performance and outputs (**Table S6**). In contrast, assuming *p*_*d*_=20% or that the vaccine protected against both infection and transmission resulted in significantly worse fits. If unrecognized mpox infections transmitted less than detected cases, a slightly smaller reduction in risk behavior (84%, 95%PI 77-92%) was needed to explain the epidemic decline compared to the best-fit model. Additionally, allowing smallpox-vaccinated MSM to also change their behavior required a higher percentage of sexual contacts (52%, 95%PI 49-54%) to be avoided to reproduce the downturn, since some of these contacts were at lower risk of infection due to prior vaccination. Overall, the other sensitivity tests showed no substantial changes in results and none improved the fit compared to the best-fit model.

## DISCUSSION

According to our analysis, a substantial change in behavior in almost half the MSM in the Paris region was the key factor in curbing the 2022 mpox epidemic and triggering its decline. These findings agree with post-outbreak behavioral survey data. In contrast, our model indicated that implemented PEP vaccination or infection-induced immunity were insufficient to explain the early downturn of the outbreak.

The reduction in risk behavior was likely due to heightened community awareness about mpox, as indicated by the shorter onset-to-testing period over time pointing to better disease recognition and treatment seeking. Our model identified June 15, 2022 as the onset for the progressive reduction in sexual contacts, in line with a wide-reaching communication campaign by authorities targeting the MSM community^15^. Without risk reduction, our analysis showed that the epidemic would have peaked in late July with three times more cases. About half the MSM were predicted to have reduced their sexual contacts in response to the outbreak— consistent with behavioral changes reported in 46% of MSM in the Paris region and in 50% of Western European MSM^7^. The model suggested a 90% reduction in sexual contacts by MSM, which may have resulted from several sources: a reduction in the number of partnerships^7^; the adoption of safer sex practices reducing transmission risk per contact– the WHO survey reported that 54% of respondents avoided drug use in sexual settings^7^, and modeling results estimated 61% to 87% drop in sexual transmission in Italy^14^; or even being away– 21% of French mpox cases having traveled abroad in June or July 2022, for example^31^. We found no evidence that change in behavior was linked to high sexual activity or recent exposure to diagnosed mpox cases. Indeed, the observed proportion of MSM required to change behavior under these hypotheses did not match that reported in the ERAS survey, especially among lower-risk individuals. ERAS data also showed no significant correlation between respondents’ levels of sexual activity and their reduction in sexual partnerships during the 2022 mpox outbreak, further confirming that the adoption of risk-reducing behaviors was widespread throughout the community.

Surveillance data from France and other affected countries showed a rapid, explosive surge in cases early in the outbreak^2^. This has been reported to be driven by the heavy-tailed distribution of sexual partnerships among MSM, where a small number of individuals have disproportionately many partners, and fuel the outbreak fast acceleration^2,3,32^. Using networks integrating sexual behavioral data from surveys reproduced the observed overdispersion in transmission. We also noted that the proportion of highly active MSM among cases was larger in the growing phase than the receding phase of the epidemic (6% vs. 4%), paralleling the trends at the national level^31^ and in line with hierarchical outbreak spread^33^. While our analysis showed that about half of MSM reduced sexual contacts, we also found that reducing contacts in the most sexually active would have required only 19% of MSM reducing risk behavior to yield the same impact on the outbreak. This suggests that prioritizing high-risk groups should be favored, especially when vaccine supplies are limited. The large spontaneous change in behavior may not be sustainable in the long term, beyond our study period, and individuals who initially altered their behavior may eventually return to their previous activity levels. However, WHO data shows that 23% of MSM in Western Europe were still reducing their sexual activity one year later^7^, suggesting a lasting impact from the 2022 epidemic wave.

We found no measurable population-level impact from PEP vaccination. This was likely due to the limited availability of doses in the early phase of the outbreak and the challenges in identifying, tracking, and vaccinating exposed individuals within the optimal delay since exposure. Contacts could not be documented systematically for all cases and most vaccinated contacts (83%) received the vaccine after the recommended 4-day delay from exposure^34^. After the decline, PrEP vaccination mitigated the epidemic over the long term, ensuring a sustained control of the outbreak^5,8,12,35^.

International reports on the mechanisms responsible for the 2022 mpox epidemic decline showed different results. Our findings differ markedly from previous work suggesting that the accumulation of immunity among highly active MSM drove the observed epidemic fadeout globally^4^. Here, we found that only 17% of highly active MSM were predicted to be infected before the epidemic peak, although they contributed a disproportionate share to infected cases (6% of cases despite representing only 0.4% of the population). For saturation to explain the course of the epidemic, our model required that about half of the entire MSM community would have been infected by summer 2022, corresponding only to 4% of cases being detected. While detection probabilities during the 2022 mpox outbreak remain undetermined in France, estimates from similar countries^14,24,36^ range between 60% and 80%. We used a 60% detection probability^24^ as a reference value and found that increasing it to 80% produced similar results and model fit. Lowering the detection probability to 20% in our model led to a significantly worse fit of the observed wave.

Behavioral adaptations are consistent with results from two modeling studies on the UK outbreak^8,12^. They estimated that the epidemic decline resulted from 45% reduction in the sexual partner rate in one model^12^, and from a 40% decrease in the transmission rate in another^8^. These estimates are close to the 39% reduction in sexual contacts resulting from our best-fit, although the second study also acknowledges an important role of infection-induced immunity. In the Netherlands, even if the epidemiological situation (1,000 cases by August 8^37^) was similar to that in the Paris region (1,357 cases by the same date), behavioral changes were not as instrumental to the reduction in cases, given that a reduction by 15-20% was sufficient to account for the decline^5^. Two studies in the US found that PrEP vaccination and behavioral changes together prevented between 64% and 84% of cases^11,13^. However, the mpox outbreak started later in the US and peaked in mid-August^2^, whereas the PrEP vaccination campaign had begun on June 26. This gave the US ample time for post-vaccine collective immunity to significantly protect the population and contribute to the epidemic decline. Although both studies recognize the role of behavioral adaptations, it was less important than in the French context where PrEP started well after the decline.

This study presents some limitations. First, we did not model pre-symptomatic transmission^38^, similarly to other modeling studies^8,12,13^. Asymptomatic transmission was addressed by including undetected milder cases and testing lower transmissibility in our sensitivity analyses, without altering our main findings. Second, we assumed a constant detection probability over time. As a result, part of the estimated behavioral change could be due to an increased detection rate over time. However, our findings on behavioral change are supported by post-outbreak survey data and hold in the range 60% to 80% for case detection, estimated for other countries. Third, ERAS is based on a convenience-based sample recruited online and through social media, as is often the case for surveys related to MSM^39^. The findings from this survey may not fully represent the experiences of everyone in the community. Finally, our three scenarios for sexual contact reduction did not cover all possible nuances. Better documenting these aspects could be the objective of future surveys.

Our findings highlight the critical role of adaptive community behavior in successfully controlling outbreaks like mpox. This is especially relevant during periods when vaccination coverage is still ramping up^40^. It underscores the power of effective risk communication as a cornerstone of outbreak management, particularly when targeting highly vulnerable communities. In the ongoing mpox crisis^1,40^, as new waves of infection caused by clade I, vaccine distribution challenges, and transmission across various settings persist, these insights are yet more vital. Empowering communities with timely, clear, and accessible information can drive significant behavioral changes that, when combined with vaccination efforts, can substantially reduce transmission. Future public health strategies for emerging pathogens must prioritize a dual approach: fostering behavioral adaptations while accelerating and expanding vaccine access to ensure both short-term mitigation and long-term epidemic control.

## Data Availability

Data from PREVAGAY and ERAS studies can be made available to researchers upon request to Annie Velter, subject to approval of the proposed analyses and agreement to adhere to security, confidentiality, and collaborative conditions.

## ACKNOWLEDGMENTS

The authors thank Simon Cauchemez, Pascal Crépey, Shweta Bansal, and Moritz U G Kraemer for useful discussions on this study. We acknowledge the Workshop of Scientific Evolutionary Writing, 5^th^ edition (sew-workshop.org/rome_2024/), where part of this paper was written.

## AUTHORS’ CONTRIBUTIONS

VC conceived and designed the study. DM, OR, PYB, MM contributed to the methodology. DM and MM analyzed the surveillance data. OR and PYB developed the code for the generation of the MSM sexual network. DM developed the code for the inference and the modeling of the transmission dynamics, ran the simulations, and analyzed the data. AV coordinated the ERAS survey, contributing to its design, online publishing, and data management. AV and DM analyzed the ERAS survey data. All authors interpreted the results. DM and VC wrote the initial manuscript draft. All authors edited and approved the final version of the Article.

## CONFLICT OF INTEREST

The authors declare no conflict of interest.

## FUNDING

This study was partially funded by: ANRS-MIE grant MPX-SPREAD (ANRS0292); ANR grant DATAREDUX (ANR-19-CE46-0008-03); EU Horizon 2020 grant MOOD (H2020-874850); EU Horizon Europe grants VERDI (101045989) and ESCAPE (101095619).

## Footnotes

* Men-who-have-sex-with-men (MSM) refers to all men who engage in sexual relations with other men. The words “men” and “sex” are interpreted differently in diverse cultures and societies and by the individuals involved. Therefore, the term encompasses the large variety of settings and contexts in which male-to-male sex takes place, regardless of multiple motivations for engaging in sex, self-determined sexual and gender identities and various identifications with any particular community or social group (Consolidated guidelines on HIV, viral hepatitis and STI prevention, diagnosis, treatment and care for key populations. Geneva: World Health Organization; 2022 (https://apps.who.int/iris/handle/10665/360601, accessed September 1, 2023).).

## Notes

### Competing Interest Statement

The authors have declared no competing interest.

### Author Declarations

All surveillance data were anonymised before use. Surveillance was considered as non interventional research only requiring the non opposition of the patient (article L1211 of the French public health code). The Comite de protection des personnes Ile de France IX (n 2014 A01605 42) authorized the PREVAGAY study. Inserm's Ethics Evaluation Committee (IRB00003888 avis n 23 989) approved the ERAS study.

## REFERENCES

1. WHO. WHO Director-General declares mpox outbreak a public health emergency of international concern. https://www.who.int/news/item/14-08-2024-who-director-general-declares-mpox-outbreak-a-public-health-emergency-of-international-concern (accessed 10 September 2024).

2. Paredes, M. I. et al. Underdetected dispersal and extensive local transmission drove the 2022 mpox epidemic. Cell 187, 1374–1386.e13 (2024).

3. Endo, A. et al. Heavy-tailed sexual contact networks and monkeypox epidemiology in the global outbreak, 2022. Science 378, 90–94 (2022).

4. Murayama, H. et al. Accumulation of Immunity in Heavy-Tailed Sexual Contact Networks Shapes Mpox Outbreak Sizes. J. Infect. Dis. 229, 59–63 (2024).

5. Xiridou, M. et al. The Fading of the Mpox Outbreak Among Men Who Have Sex With Men: A Mathematical Modelling Study. J. Infect. Dis. 230, e121–e130 (2023).

6. Delaney, K. P. et al. Strategies Adopted by Gay, Bisexual, and Other Men Who Have Sex with Men to Prevent Monkeypox virus Transmission — United States, August 2022. MMWR Morb. Mortal. Wkly. Rep. 71, 1126–1130 (2022).

7. Prochazka, M. et al. Temporary adaptations to sexual behaviour during the mpox outbreak in 23 countries in Europe and the Americas: findings from a retrospective cross-sectional online survey. Lancet Infect. Dis. In press (2024) doi:10.1016/S1473-3099(24)00531-0.

8. Brand, S. P. C. et al. The role of vaccination and public awareness in forecasts of Mpox incidence in the United Kingdom. Nat. Commun. 14, 4100 (2023).

9. Van Dijck, C., Hens, N., Kenyon, C. & Tsoumanis, A. The Roles of Unrecognized Mpox Cases, Contact Isolation and Vaccination in Determining Epidemic Size in Belgium: A Modeling Study. Clin. Infect. Dis. 76, e1421–e1423 (2023).

10. Milwid, R. M. et al. Exploring the dynamics of the 2022 mpox outbreak in Canada. J. Med. Virol. 95, e29256 (2023).

11. Clay, P. A. et al. Modelling the impact of vaccination and sexual behaviour adaptations on mpox cases in the USA during the 2022 outbreak. Sex. Transm. Infect. 100, 70–76 (2023).

12. Zhang, X.-S. et al. Transmission dynamics and effect of control measures on the 2022 outbreak of mpox among gay, bisexual, and other men who have sex with men in England: a mathematical modelling study. Lancet Infect. Dis. 24, 65–74 (2024).

13. Lin, Y.-C., Wen, T.-H., Shih, W.-L., Vermund, S. H. & Fang, C.-T. Impact of vaccination and high-risk group awareness on the mpox epidemic in the United States, 2022–2023: a modelling study. eClinicalMedicine 68, 102407 (2024).

14. Guzzetta, G. et al. The decline of the 2022 Italian mpox epidemic: Role of behavior changes and control strategies. Nat. Commun. 15, 2283 (2024).

15. Ministère de la santé et de l’accès aux soins. Monkeypox : le point sur le virus. https://sante.gouv.fr/soins-et-maladies/maladies/maladies-infectieuses/monkeypox/cas-groupes-d-infection-par-le-virus-monkeypox (accessed 30 March 2023).

16. Santé Publique France. Cas de mpox (monkeypox) détectés en France. https://www.santepubliquefrance.fr/maladies-et-traumatismes/maladies-transmissibles-de-l-animal-a-l-homme/monkeypox/donnees/#tabs (accessed 23 October 2022).

17. Haute Autorité de Santé. Avis n°2022.0034/SESPEV du 20 mai 2022 du collège de la Haute Autorité de santé relatif à la vaccination contre Monkeypox. https://www.has-sante.fr/jcms/p_3340378/fr/avis-n2022-0034/sespev-du-20-mai-2022-du-college-de-la-haute-autorite-de-sante-relatif-a-la-vaccination-contre-monkeypox (accessed 30 March 2023).

18. Haute Autorité de Santé. Avis n°2022.0039/AC/SESPEV du 7 juillet 2022 du collège de la Haute Autorité de santé relatif à la vaccination contre le virus Monkeypox en préexposition des personnes à haut risque d’exposition. https://www.has-sante.fr/jcms/p_3351308/fr/avis-n2022-0039/ac/sespev-du-7-juillet-2022-du-college-de-la-haute-autorite-de-sante-relatif-a-la-vaccination-contre-le-virus-monkeypox-en-preexposition-des-personnes-a-haut-risque-d-exposition (accessed 30 March 2023).

19. Santé Publique France. Cas de Mpox en Europe, définitions et conduite à tenir. 6 https://www.santepubliquefrance.fr/media/files/maladies-a-declaration-obligatoire/definition-de-cas-cat-monkeypox (accessed 20 April 2023).

20. Robineau, O., Velter, A., Barin, F. & Boëlle, P.-Y. HIV transmission and pre-exposure prophylaxis in a high risk MSM population: A simulation study of location-based selection of sexual partners. PLOS ONE 12, e0189002 (2017).

21. Sommen, C. et al. Time location sampling in men who have sex with men in the HIV context: the importance of taking into account sampling weights and frequency of venue attendance. Epidemiol. Infect. 146, 913–919 (2018).

22. Velter Annie, Duchesne Lucie, Lydié Nathalie. EVOLUTION DES COMPORTEMENTS DE PRÉVENTION CHEZ LES HOMMES AYANT DES RAPPORTS SEXUELS AVEC DES HOMMES EN FRANCE - ENQUÊTES RAPPORT AU SEXE 2017 ET 2019. https://www.santepubliquefrance.fr/maladies-et-traumatismes/infections-sexuellement-transmissibles/vih-sida/documents/communication-congres/evolution-des-comportements-de-prevention-chez-les-hommes-ayant-des-rapports-sexuels-avec-des-hommes-en-france-enquetes-rapport-au-sexe-2017-et-2019 (accessed 13 July 2023).

23. Velter, A., Karen, C., Girard, G., Roux, P. & Mercier, A. HIV Pre-exposure prophylaxis among men who have sex with men responding to the Rapport au Sexe 2023 survey: Who is eligible? Who are the users? https://beh.santepubliquefrance.fr/beh/2023/24-25/2023_24-25_5.html (accessed 28 May 2024).

24. Borges, V. et al. Viral genetic clustering and transmission dynamics of the 2022 mpox outbreak in Portugal. Nat. Med. 29, 2509–2517 (2023).

25. Colombe, S. et al. Effectiveness of historical smallpox vaccination against mpox clade II in men in Denmark, France, the Netherlands and Spain, 2022. Eurosurveillance 29, 2400139 (2024).

26. Fine, P. E. M., Jezek, Z., Grab, B. & Dixon, H. The Transmission Potential of Monkeypox Virus in Human Populations. Int. J. Epidemiol. 17, 643–650 (1988).

27. Payne, A. B. et al. Reduced Risk for Mpox After Receipt of 1 or 2 Doses of JYNNEOS Vaccine Compared with Risk Among Unvaccinated Persons — 43 U.S. Jurisdictions, July 31–October 1, 2022. MMWR Morb. Mortal. Wkly. Rep. 71, 1560–1564 (2022).

28. Montero Morales, L. et al. Post-exposure vaccine effectiveness and contact management in the mpox outbreak, Madrid, Spain, May to August 2022. Eurosurveillance 28, 2200883 (2023).

29. Bertran, M. et al. Effectiveness of one dose of MVA–BN smallpox vaccine against mpox in England using the case-coverage method: an observational study. Lancet Infect. Dis. 23, S1473309923000579 (2023).

30. Christodoulidou, M. M. & Mabbott, N. A. Efficacy of smallpox vaccines against Mpox infections in humans. Immunother. Adv. 3, ltad020 (2023).

31. Krug, C. et al. Mpox outbreak in France: epidemiological characteristics and sexual behaviour of cases aged 15 years or older, 2022. Eurosurveillance 28, 2200923 (2023).

32. Pastor-Satorras, R. & Vespignani, A. Epidemic Spreading in Scale-Free Networks. Phys. Rev. Lett. 86, 3200–3203 (2001).

33. Barthélemy, M., Barrat, A., Pastor-Satorras, R. & Vespignani, A. Velocity and Hierarchical Spread of Epidemic Outbreaks in Scale-Free Networks. Phys. Rev. Lett. 92, 178701 (2004).

34. Santé Publique France. Bilan du suivi des contacts à risque des cas probables et confirmés d’infection par le virus Monkeypox, mai - juillet 2022. https://www.santepubliquefrance.fr/maladies-et-traumatismes/maladies-transmissibles-de-l-animal-a-l-homme/monkeypox/documents/rapport-synthese/bilan-du-suivi-des-contacts-a-risque-des-cas-probables-et-confirmes-d-infection-par-le-virus-monkeypox-mai-juillet-2022 (accessed 2 November 2023).

35. Ghosn, J. et al. Impact of vaccination with third generation modified vaccinia Ankara and sexual behaviour on mpox incidence in men who have sex with men: analysis among participants of the ANRS-174 DOXYVAC trial. Lancet Reg. Health - Eur. 45, 101020 (2024).

36. Polk, C. et al. Evaluation of a health system’s implementation of a monkeypox care model under the RE-AIM framework. Ther. Adv. Infect. Dis. 10, 204993612311584 (2023).

37. Van Ewijk, C. E. et al. Mpox outbreak in the Netherlands, 2022: public health response, characteristics of the first 1,000 cases and protection of the first-generation smallpox vaccine. Eurosurveillance 28, 2200772 (2023).

38. Ward, T., Christie, R., Paton, R. S., Cumming, F. & Overton, C. E. Transmission dynamics of monkeypox in the United Kingdom: contact tracing study. BMJ 379, e073153 (2022).

39. Beyrer, C. et al. Global epidemiology of HIV infection in men who have sex with men. The Lancet 380, 367–377 (2012).

40. Ndembi, N. & Abdool Karim, S. S. Africa aims to avert an mpox pandemic. Science 386, 7 (2024).

